# First-in-clinical application of a time-gated diffuse correlation spectroscopy system at 1064nm using superconducting nanowire single photon detectors

**DOI:** 10.1101/2021.11.11.21266071

**Authors:** Chien-Sing Poon, Dharminder S. Langri, Benjamin Rinehart, Timothy M. Rambo, Aaron J. Miller, Brandon Foreman, Ulas Sunar

## Abstract

Recently proposed time-gated DCS (TG-DCS) has significant advantages compared to conventional CW-DCS, but it is still in an early stage and clinical capability has yet to be established. The main challenge for TG-DCS is the lower SNR when gating for the deeper travelling late photons. Longer wavelengths, such as 1064nm have a smaller effective attenuation coefficient and a higher power threshold in humans, which significantly increases the SNR. Here, we demonstrate the clinical utility of TG-DCS at 1064nm in a case study on a patient with severe traumatic brain injury admitted to the neuroscience intensive care unit (NSICU). We showed a significant correlation between TG-DCS early (ρ = 0.67) and late (ρ = 0.76) gated against invasive thermal diffusion flowmetry. We also analyzed TG-DCS at high temporal resolution (50 Hz) to elucidate pulsatile flow data. Overall, this study demonstrates the first clinical translation capability of the TG-DCS system at 1064nm using superconducting nanowire single photon detector.

## Introduction

Traumatic brain injury (TBI) is one of the leading causes of death and disability, contributing up to 30% of all injuries-related deaths in the United States and resulting in long-term disability with an annual cost of $80 billion [1]. With more than 2.5 million emergency department visits, 288,000 hospitalizations, and 57,000 deaths related to TBI, it is a significant public health burden. From 2006 to 2014, the number of emergency department visits, hospitalizations, and deaths related to TBI increased by 53%, while in 2014 alone an average of 155 people in the United States died each day from TBI-related injuries.

Numerous events can cause TBI, including mechanical falls, traffic accidents, military combat injuries, and sports injuries, among others. The diagnosis and evaluation of brain injury is crucial in composing a treatment plan for complications associated with brain trauma. These complications include changing levels of consciousness, memory disturbances, confusion, deficits in orientation, new-onset or worsening seizures, auditory, visual, and emotional complications, and death. However, current diagnostic methods for TBI-related complications are limited to self-reporting symptoms, functional assessments such as the Glasgow Coma Scale, speech tests, language tests, cognitive assessments, and neurophysiological tests.

Several imaging technologies are available for clinical use, including computed tomography (CT) and magnetic resonance imaging (MRI). Although these are non-invasive, brain imaging provides only single snapshots in time and may not capture evolving injuries or their complications. In the most severe brain injuries, complications due to TBI – termed secondary brain injuries – can occur quickly and cause significant morbidity or mortality. These patients are cared for in intensive care units (ICUs) focused on monitoring the brain for secondary brain injuries. Patients may not be able to report complications and imaging tests are not suitable for the continuous bedside monitoring needed in this setting [2]. Rather, continuous monitoring in most ICUs is limited to measurements of cardiopulmonary function such as cardiac telemetry or arterial blood pressure monitoring. In selected patients, invasive probes are recommended to measure intracranial pressure (ICP) in selected patients according to the Brain Trauma Foundation Guidelines [1], [3]. In some centers, multimodality monitoring (MMM) is used that incorporates not only ICP but also measurements of brain tissue oxygen tension (PbtO2) or regional cerebral blood flow (rCBF). rCBF can be measured from brain tissue directly using a thermal diffusion flowmetry probe (TDF) (Hemedex, Inc; Cambridge, MA, USA) [4]. However, these methods are invasive, require drilling a burr hole through the skull, which increases the risk of hemorrhage and infection. Furthermore, TDF only probes a small volume and suffers from data loss due to automatic recalibration every 30 min.

Laser Doppler flowmetry (LDF) shares a similar issue with TDF in that it measures CBF only from a very small volume of tissue [5]. Transcranial Doppler ultrasound (TCD) is more sensitive to larger vessels, but it is limited by skull thickness, and it is difficult to use for longitudinal monitoring due to the need for a stable orientation of the ultrasound probe [6]. Continuous EEG (cEEG) can detect seizures, periodic discharges, and spreading depolarizations leading to secondary brain ischemia and may contain relevant prognostic information in patients after moderate or severe TBI [7]–[10]. Currently, the international neurocritical care community recommends cEEG monitoring in patients after TBI. Characteristic changes in EEG signals have been recognized in response to brain ischemia in correlation with CBF and oxygen metabolism [11], [12]. When the CBF is compromised, the metabolic and electrical activity of cortical neurons is affected, leading to changes in the frequency content of EEG [12]. The disadvantage of EEG is its acquisition paradigm, which produces a high-amplitude mixed frequency that makes far-field event detection difficult and the need for interpretation. Furthermore, EEG exhibits low signal-to-noise ratio (SNR) because of placement on the scalp far from the source of the signal, artifacts common in the ICU setting, and the effects of interventions, such as sedation. Non-invasive EEG therefore cannot always be relied on to inform of evolving secondary brain injury associated with TBI. Thus, there is need for continuous, non-invasive monitoring at cerebral blood flow at the bedside in the ICU setting.

Diffuse optical technologies enable continuous non-invasive measurements of the hemodynamics within the brain microvasculature [6], [13]. Near-infrared spectroscopy (NIRS) can determine changes in cerebral oxygenation, but cannot provide a direct measure of CBF [6], [13]. Furthermore, oxygen saturation alone may not be sensitive enough to detect secondary brain injury at the later time points of the insult, as oxygen consumption and delivery may reach equilibrium after acute transient alterations [6], [13], [14]. Significant work has been done on the monitoring of cerebral oxygenation with NIRS [15], [16]. For example, Robertson et al. showed a ∼90% sensitivity for the detection of secondary hematomas [17], and Kampfl et al. showed that patients with an ICP of >25 mmHg exhibited significantly reduced NIRS parameters than those with an ICP <25 mmHg [18]. Yet, there are limitations in NIRS monitoring [1,2]. Busch et al. showed that the NIRS approach was limited in detecting cerebral hypoxia in patients with acute brain injury [14]. Leal-Noval et al. found that NIRS was reliably sensitive to detect relatively severe cerebral hypoxia (PbtO2 < 12 mm Hg), but showed a weak correlation between measurements of invasive PbtO2 and noninvasive tissue oxygenation measured by NIRS [19]. Overall, NIRS studies have indicated that there was no strong evidence of a significant correlation with invasive monitoring modalities [20]. Almost all of these studies used commercial NIRS systems to provide information related to changes in oxygenation rather than absolute values, and these changes did not correlate well with the clinical outcome [21]. These systems work in continuous-wave (CW) mode and cannot effectively separate the scalp signal from the deeper brain signal. Regression analysis and adding and adding short source-detector separation [6], [22]–[26] can provide improvements for separating superficial vs. deeper brain signals, but most of the commercial NIRS instruments still do not have this type of optode configurations.

A more recently developed diffuse optical technique, diffuse correlation spectroscopy (DCS), probes speckle fluctuations related to blood flow [6], [13], [27]–[30]. It is ideal for high-risk populations, as it can be used as a portable, non-invasive, continuous bedside monitor for CBF [6], [14], [31]–[35]. There is a much higher contrast between cerebral blood flow vs. scalp blood flow [35]. Thus, we expect that blood flow measurements may provide additional benefits for TBI monitoring [14], [35]. In fact, DCS has been shown to have a higher sensitivity to the brain compared to NIRS [36]. DCS has shown promise for noninvasive, continuous measurements of CBF in animals and humans [30], [37]–[39], and recently, in neuro-ICU settings [6], [14], [31]–[35]. It has been demonstrated that DCS measurements agree with clinically-established modalities [22,25,35]. However, most DCS operates in the continuous wave (CW) domain. There exist several crucial limitations of CW-DCS regarding (1) the ability to obtain absolute blood flow (2) its sensitivity to deeper target tissue, (3) the occupation of limited space on the forehead, (4) its tissue probing depth, and (5) the SNR within deeper tissue. The work by Baker et al. is highly significant in this aspect since it directly tackles the limitations of CW-DCS approach by applying more realistic two-layer (scalp and brain) model in their data analysis to separate scalp and brain signals [26]. The contamination from the scalp signal in CW-DCS can be reduced by regression analysis and using short source-detector optode configurations [6], [40], [41].

An elegant way of separating path-length resolved blood flow is a recent development of time-gated diffuse correlation spectroscopy (TG-DCS) [42], [43], [52]–[60], [44]–[51], which can also quantify (1) absolute blood flow by obtaining the absorption (µ_a_) and reduced scattering (µ_s_’) parameters, which can be used as a priori information during the data analysis for blood flow related parameter quantification. The time-gating aspect allows selecting deeper photons for (2) greater brain blood flow sensitivity without the need for long source-detector separations. This reduces the size of the probe for (3) greater compactness. Short-wave infrared (SWIR) wavelengths such as1064 nm has a low effective attenuation in brain tissue [44], [47], [53], [54], which (4) increases the tissue probing depth, and allows for a higher maximum permissible laser power (ANSI Z136.1 [61]) to (5) increase the SNR. Very recently we demonstrated the clinical feasibility of using a new generation photon counting detector, superconducting nanowire single photon detector (SNSPD) for CW-DCS with improved SNR at 785 nm [62]. In this paper, we show the clinical feasibility of a TG-DCS by using SNSPD, which enabled us to use of a SWIR wavelength at 1064 nm in a patient with severe TBI in an ICU setting. We compared our TG-DCS results with the invasive TDF monitor. Additionally, we sought to provide evidence that the sampling frequency of TG-DCS is sufficient to resolve pulsatile blood flow by correlating it with a commercial heart rate monitor.

## Material and methods

### Study Design and Patient Details

A clinical prototype of the TG-DCS system was built and tested in a patient with TBI admitted to the neuroscience intensive care unit (NSICU) at the University of Cincinnati Medical Center after informed consent from a legally authorized representative as approved by the Institutional Review Board (IRB) of the University of Cincinnati. This case involves a man in his 20s, who was involved in a vehicular accident. He was helmeted but upon admission to the Emergency Department, he was found to have depressed level of arousal with a Glasgow Coma Scale score of 7T. Both pupils were reactive to light. A non-contrast head CT demonstrated a left subdural hematoma with left-to-right midline shift (**Fig. 1A**). Hypertonic saline was administered, and he was taken urgently to the operating room, where he underwent left decompressive hemicraniectomy and evacuation of the clot. His additional injuries included bilateral pulmonary contusions. Commercially available, FDA-approved, invasive multimodality intracranial monitoring (MMM) probes were placed within non-dominant frontal subcortical white matter using a burr hole created at the Kocher’s point approximately between scalp EEG channels F4 and C4 (according to measurements using the International 10-20 system; **Fig. 1D**). One of these probes was a thermal diffusion flowmetry device (Bowman Perfusion Monitor; Hemedex, Inc; Cambridge, MA) placed to directly measure rCBF in conjunction with other monitored modalities, including ICP, PbtO2, cerebral microdialysis, and electrocorticography as previously described [63].

**Figure 1.**
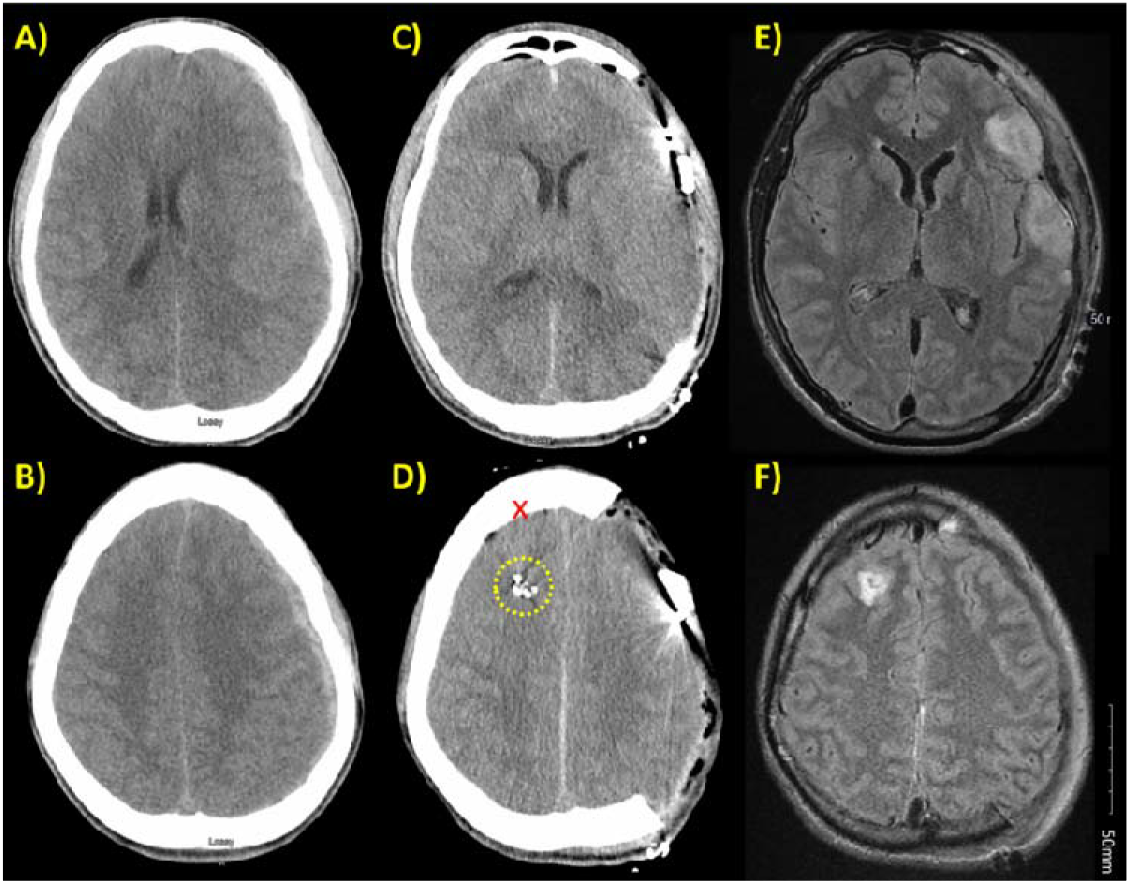
Computed Tomography (CT) head imaging. (**A, B**) Pre-operative imaging at admission demonstrated left to right midline shift. (**C, D**) Postoperative imaging after decompressive hemicraniectomy. Multimodal Intracranial Monitoring (MMM) probes were placed in the contralateral hemisphere approximately 2.5cm below the inner table of the skull within the subcortical white matter. The location of the MMM is highlighted with a yellow circle. The approximate location of the time-gated diffusion correlation spectroscopy (TG-DCS) probe is marked with an ‘x’. (**E, F**) Magnetic resonance imaging (MRI) on the head on post-trauma day 4 with the MMM removed.

On post-trauma day 3, the portable TG-DCS system was brought to the NSICU. The system was placed in the patient’s room and a total of 1 h and 20 min of data were collected along with simultaneously-recorded MMM measurements. During the duration of the TG-DCS measurement, three events related to CBF occurred: (1) propofol administration was stopped, (2) a neurological examination was performed, and (3) administration of propofol was resumed as shown in **Fig. 2**.

**Figure 2.**
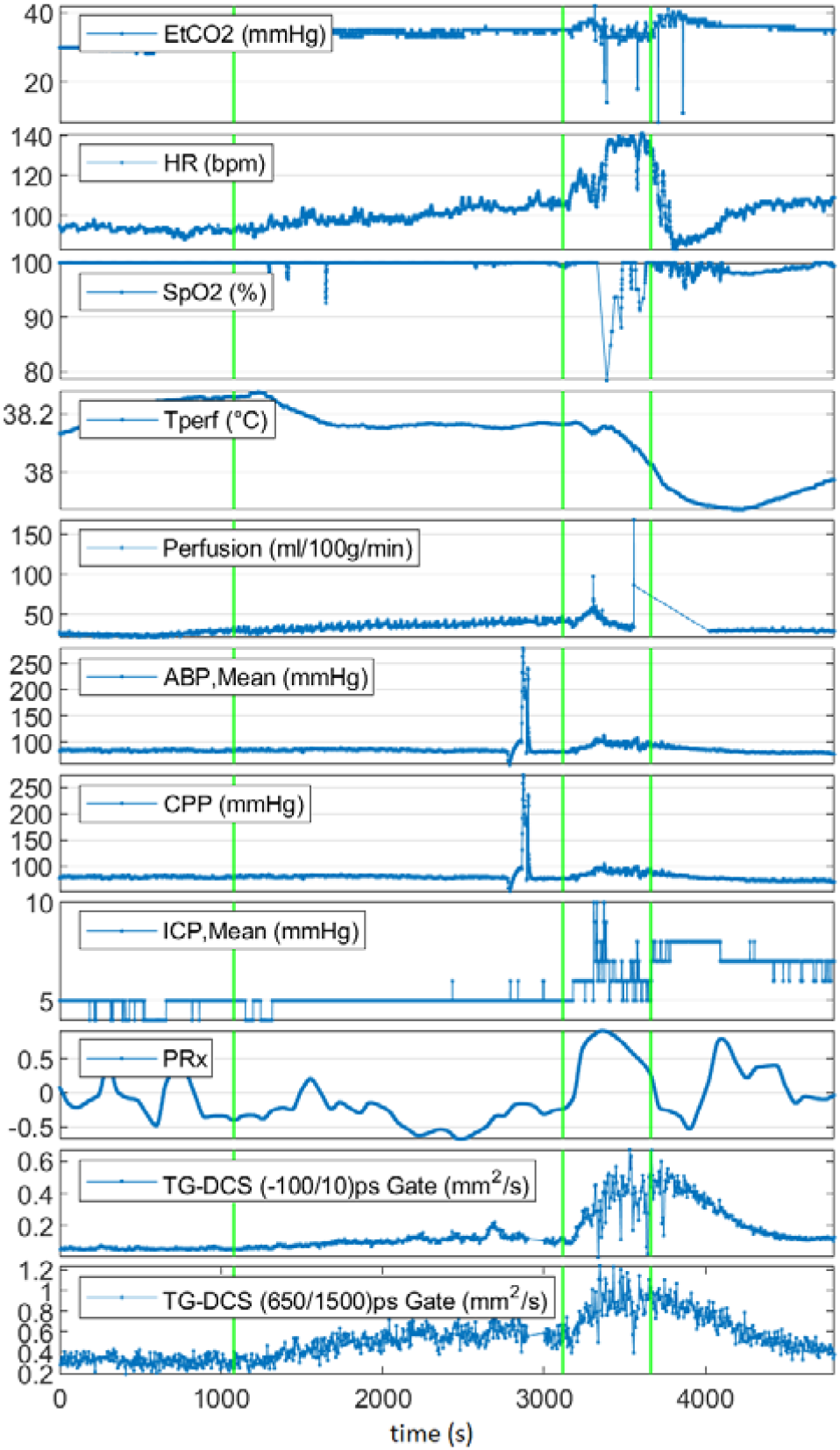
Data from the invasive multimodal monitor probes are plotted with the time-gated DCS (TG-DCS) early (−100ps from the peak, with a width of 10ps) and late (650ps from the peak with 800 ps width) blood flow index. The 3 green line represents (1) the time that sedation is stopped, 2) the neurological examination is performed, and (3) the sedation is restarted.

### Optical Technique

A portable prototype TG-DCS was developed and placed in the patient’s room (**Fig. 3A**) during the measurement period. The optical imaging probe was placed just behind scalp EEG electrode F4 and above the F8 electrode. The probe consisted of a 1 mm source fiber and a few-mode fiber bundle (4 fibers) at 15 mm source detector separation. The fibers were mounted onto a 5 mm right-angled prism and housed in a custom printed flexible 3D probe (**Fig. 3B**). The probe was ergonomically designed to allow for easy placement and designed to minimize area of scalp needed, with a width just slightly larger than a US quarter dollar (**Fig. 3C**). The probe was sanitized with an alcohol swab and held in position with a Tegaderm™ film (3M; Saint Paul, MN). All four channels were connected to the SNSPD to collect the reflected photons at 1064 nm. Each of the active channels was connected to the 4-channel time-correlated single-photon counter (TCSPC) (HydraHarp400, Picoquant Inc, Berlin, Germany). The laser source consists of a 1064nm seed laser, operated at a repetition rate of 80MHz with a pulse width of 250ps (QLD-106P, QDLaser Inc, Kanagawa, Japan). The seed laser was amplified with a 1064nm optical amplifier (Mantaray-Amp-1064, Cybel LLC, Bethlehem, PA) and was fiber-coupled to a 1mm multimode fiber that was mounted onto the 3D printed probe. The beam on the face of the prism was expanded to an approximately 4.5 mm diameter spot and the average power was set below ANSI standards at 150 mW (∼943 mW/cm^2^). Each channel sampled at approximately 1 × 10^7^ cps.

**Figure 3.**
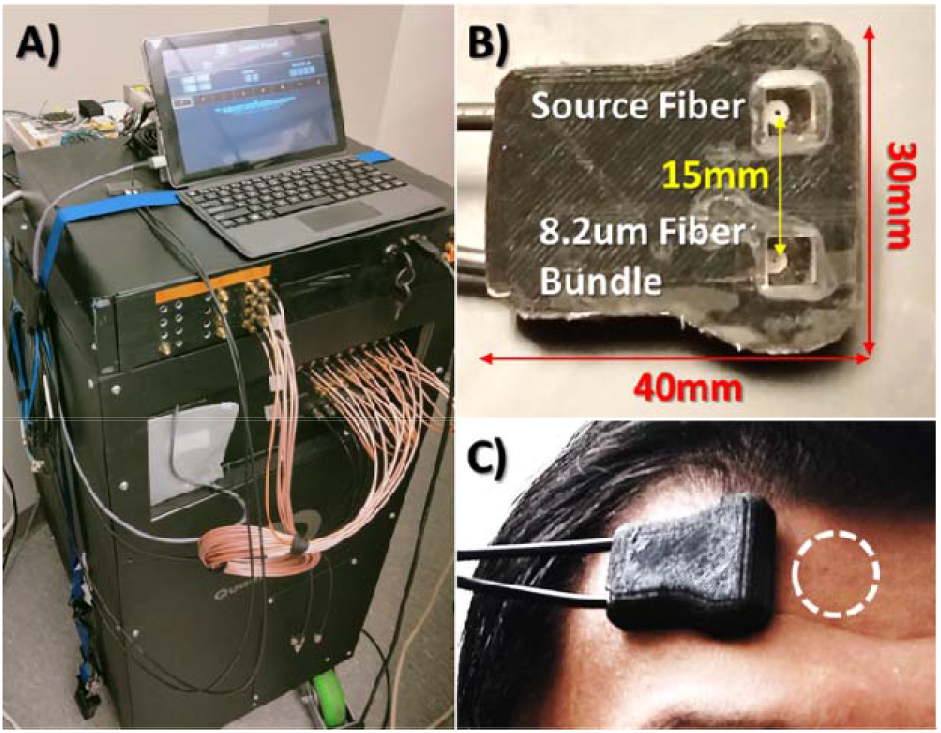
**(A)** The clinical TDDCS system that was used in the NICU. **(B)** Image of the ergonomic 3D printed probe used on the patient. **(C)** An example of probe placement on the forehead with the approximate size of a US quarter dollar in white as a comparison of the size.

### Data Analysis

Time-tagged data was collected in 8 × 10 minute bins, representing a total of ∼140GB of data at ∼1.75GB/minute of monitoring. The data was loaded into a custom MATLAB routine on a supercomputer node with 1.5TB of RAM[64]. The instrument response function (IRF) was obtained by placing a thin scattering layer (Teflon Tape, 3M, Saint Paul, MN) between the source fiber and one of the detector fibers for an acquisition time of 30 s. The full width half-maximum (FWHM) of the pulse was measured to be ∼220ps (**Fig. 4A**). Two gates were selected: early gate (EG) (−100ps from the peak with a width of 10ps) and late gate (LG) (+650ps from the peak with a width of 1ns) (**Fig. 4B**). The EG and LG data was autocorrelated from 5e-7s to 1e-5s and integrated for 5s (**Fig. 4C**). The optical properties of µs’: 0.83mm^-1^ and µa: 0.018mm^-1^. Were used in the g2 model in the time domain [38,39,46] to fit the data for the absolute blood flow index (BFi) (**Fig. 4D**).

**Figure 4.**
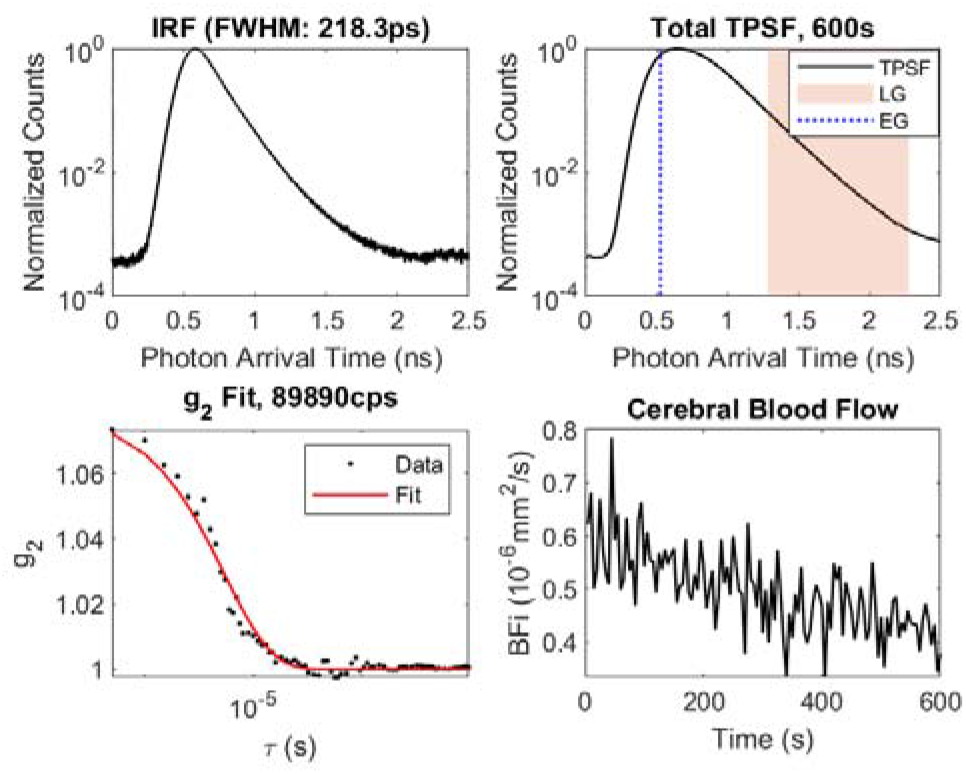
**(A)** Instrument response function (IRF) obtained from the laser. **(B)** Representative TPSF curve of the patient with the early (EG) and late (LG) gates highlighted. The EG opens -100ps from the peak with a width of 10ps (dashed line), while the LG opens +650ps from the peak with a width of 1500ps (red overlay) **(C)** Representative g_2_ curve of the patient that is fitted with the time-gated g_2_ model. **(D)** The absolute BFi obtained from the fit.

## Results and discussion

We obtained a Spearman correlation coefficient (ρ) between TD-DCS BFi and TDF rCBF of 0.67 (EG) and 0.76 (LG) (p < 0.001 for both; **Fig. 5A, B**). Examining the time-series of the TG-DCS BFi vs. the TDF rCBF signal, we observed a greater increase in LG versus EG after the first green line (sedation turned off). At the second green line, a neurological examination was performed that included painful stimulation to test the level of arousal and motor functioning. A sharp increase in CBF was observed in both EG and LG. The magnitude of the change in rCBF triggered a calibration cycle of the TDF device, which is expected behavior designed to guarantee the accuracy of its measurements (see the data gap beginning ∼ 3500 s). The third green line marks the restart of sedation, which caused a decrease in both TG-DCS BFi and TDF rCBF.

**Figure 5.**
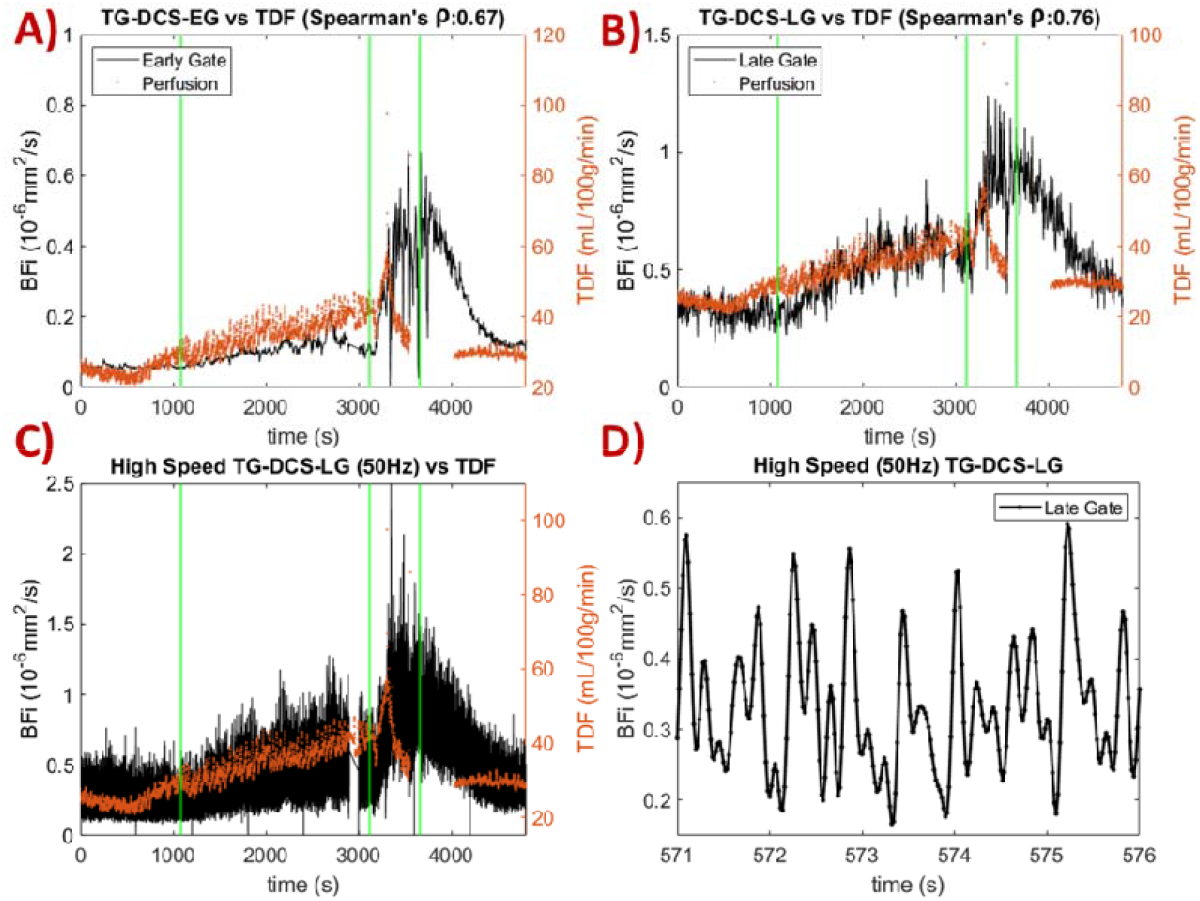
**(A)** Early-gate blood flow index (EG-BFi) **(B)** LG-BFi vs. invasive TDF CBF. Spearman’s correlation, R, of 0.67 and 0.76 was obtained, respectively. **(C)** The 50Hz late-gate BFi shows greater fluctuations in BFi due to pulsatile blood flow. **(D)** A zoomed-in portion of the 50Hz BFi showing pulsatile blood flow. The 3 green lines represent annotations at which (1) sedation was stopped, (2) a neuro exam was performed, and (3) sedation was restarted.

To show the feasibility of TG-DCS at high temporal resolution (50Hz), wider gates closer to the peak of temporal point spread function (TPSF) were used to increase the signal-to-noise ratio (SNR). The opening time/gate width relative to the peak was adjusted to +200ps/1800ps. The data were correlated at a rate of 50Hz, which is sufficient to resolve heart rate up to 300 beats per minute. At higher resolution, we observed greater fluctuations in the BFi (**Fig. 5C**). When a portion of the data is zoomed in, we observed a pattern like that of the pulsatile flow, which is related to the heart rate (**Fig. 5D**).

We analyzed the BFi at 10Hz to have greater sensitivity to the superficial and deep BFi while maintaining good SNR and temporal resolution. The EG opening time/gate width relative to the peak was adjusted to -150ps/100ps while the LG opening time/gate width relative to the peak was adjusted to +400ps/1600ps. To validate that we can obtain pulsatile blood flow, a second order zero phase Butterworth filter with a passband of 0.75-3Hz (45-180bpm) was used. The signal was then normalized by first subtracting the mean then dividing by the standard deviation of itself. Next, the Welch’s method with 64 windows and 50% overlap was used to obtain the power spectral density (PSD) of the TG-DCS BFi (**Fig. 6**). We compared the frequency spectra to the frequencies of the heart rate (HR) signal, which was recorded using a bedside data aggregation system to ensure signals were time-locked (Component Neuromonitoring System; Moberg Solutions, Inc; Ambler, PA). The PSD obtained from TG-DCS was overlaid with the PSD from the HR (**Fig. 6**). A ρ of 0.89 (LG vs HR) and 0.80 (EG vs HR) (p<0.001 for both) were obtained showing a significant correlation between TG-DCS and HR signals.

**Figure 6.**
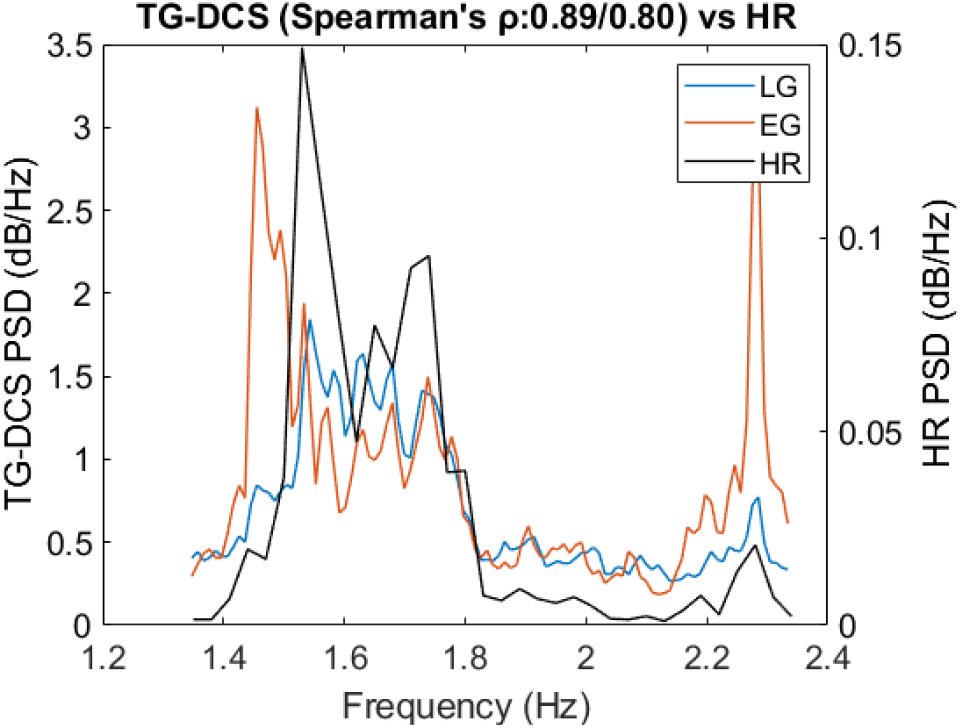
The PSD of EG and LG was compared with the heart rate (HR) obtained from a commercial instrument, showing significant correlation.

## Conclusion

This is the first reported clinical use of TG-DCS using SNSPD at 1064 nm to resolve continuous, pulsatile blood flow within brain tissue that is currently accessible only by invasive methods. Our compact probe was able to measure BFi, which correlated well with the rCBF obtained from an invasive thermal diffusion-based technique. Non-invasive measurement could provide continuous blood flow data during a period in which invasive measures were limited by the need for signal calibration. Additionally, we were able to adjust the photon gating to manipulate the temporal resolution such that pulsatile blood flow could be resolved, which was verified against the heart rate recorded using a commercial clinical device. The ability to adjust the SNR, depth, and sampling frequency provided an exceptionally flexible tool that can be customized to the patient and to resolve specific physiological indicators, such as the relationship between arterial pressure and cerebral blood flow that could ultimately be used to estimate intracranial pressure or cerebral autoregulation.

## Data Availability

All data produced in the present study are available upon reasonable request to the authors

## Acknowledgements

The authors thank the NIH funding support, R01 (NIBIB Brain Initiative, 1R01EB031759), R03 (NINDS, 5R03NS115022). Additional support was provided by the Ohio Third Frontier to the Ohio Imaging Research and Innovation Network (OIRAIN). We also thank Nick Bertone (Picoquant Inc.) for providing the demo unit of HydraHarp400 for the time domain acquisition system, and the Ohio Supercomputer Center for providing us with a supercomputer facility to perform the analysis.

